# Linking climate change to self-harm: A global study of over 200 countries from 1990 to 2020

**DOI:** 10.1101/2025.08.04.25332918

**Authors:** Duan Ni, Ian B. Hickie, Mark Howden, Ralph Nanan

## Abstract

Climate change significantly affects both environmental and human health. A recent, comprehensive global overview of its impacts on mental health and self-harming behaviours is lacking, as well as detailed insights into the effects of potential confounders, including gender, age, socioeconomic factors and their potential interactions.

Using a cutting-edge generalized additive model (GAM) framework, we analyzed multiple global datasets covering between 175 to 201 countries from 1990 to 2020. We found robust associations between self-harm incidence rates and major climate change parameters, including greenhouse gas emission and temperature change, as well as air pollutant particulate matter 2.5 (PM2.5) exposure, a critical contributor to climate change. Importantly, detailed analyses suggested that self-harm in young males had stronger links to climate change parameters than in young females, while the opposite gender associations were found later in life.

Our global analyses provide important evidence on mental health consequences of climate change, instructive for developing appropriate population-based mental health strategies and climate policies and enhancing mental health services. This will contribute to improving human and planetary health.

## Introduction

Climate change is an existing and growing challenge to both environmental and human health^1^. Alarmingly, the incidence of common mental disorders, particularly among young people, appears to be rising steeply in recent decades^2,3^. Accumulating evidence from regional cohort studies and derivate meta-analyses suggests that climate change affects mental health and wellbeing and might be linked directly to increased risk of self-harm and other suicidal ideas or behaviours^4-6^. However, a global overview of mental health impacts from different aspects of climate change is lacking. In this context, other known confounding variables for mental health, such as gender, age and socioeconomics^3^ also need to be factored in. For instance, self-harm appears to be more common in females than males^7,8^, and there seems to be an increasing prevalence of self-harm in adolescents^3^. Furthermore, there are well-established disparities in self-harm behaviours among countries or populations of different socioeconomic status^3^. Different socioeconomic status is likely to influence the capacity and access to resources to cope with climate change. Therefore, the potential interactive effects among these confounders also warrant investigations.

Harnessing a state-of-the-art generalized additive model (GAM) framework^9,10^, we systematically analyzed the associations between self-harm incidence and major climate change parameters like greenhouse gas (GHG) emission, temperature change and relative humidity, as well as air pollutant particulate matter 2.5 (PM2.5) exposure, an important contributor to climate change^11^, by country on a global scale. GHG emission, temperature change and PM2.5 exposure generally correlated with elevated self-harm incidence, but humidity conferred limited effects. Strikingly, we uncovered complex interactive effects among gender, age and socioeconomic status on the impacts of climate change on self-harm.

Together, we present to the best of our knowledge the first global overview of the detailed associations between climate change-related factors and self-harm. These findings highlight the likely mental health consequences of climate change and emphasize the urgent need to incorporate population-based mental health strategies and enhancement of mental health services in future climate policies.

## Results

From 1990, self-harm incidence rates continuously declined globally, with a reversal of this trend since 2019 (Figure 1a). In parallel, from 1990 to 2021, global gross domestic product (GDP) per capita, a close proxy of socioeconomic wealth, consistently increased (Figure 1b). The global per capita GHG emission (Figure 1c) and temperature changes relative to the baseline climatology during 1951-1980 (Figure 1d) also generally increased over the period 1990-2021. At the same time, population-weighted PM2.5 exposure varied considerably, with a decline between 1990 and 2010, an increase from 2010 to 2013, and a steep decrease between 2013-2020 (Figure 1e). Since most of these parameters are intercorrelated (Table S1), disentangling their relationships is challenging. Various confounders linked to self-harm^3,7,12^, such as age, gender, substance use and geographical factors, also add to the complexity. Hence, meaningful analyses required inclusion of high-quality data, multi-dimensional approaches, and the application of advanced mathematical tools.

**Figure 1.**
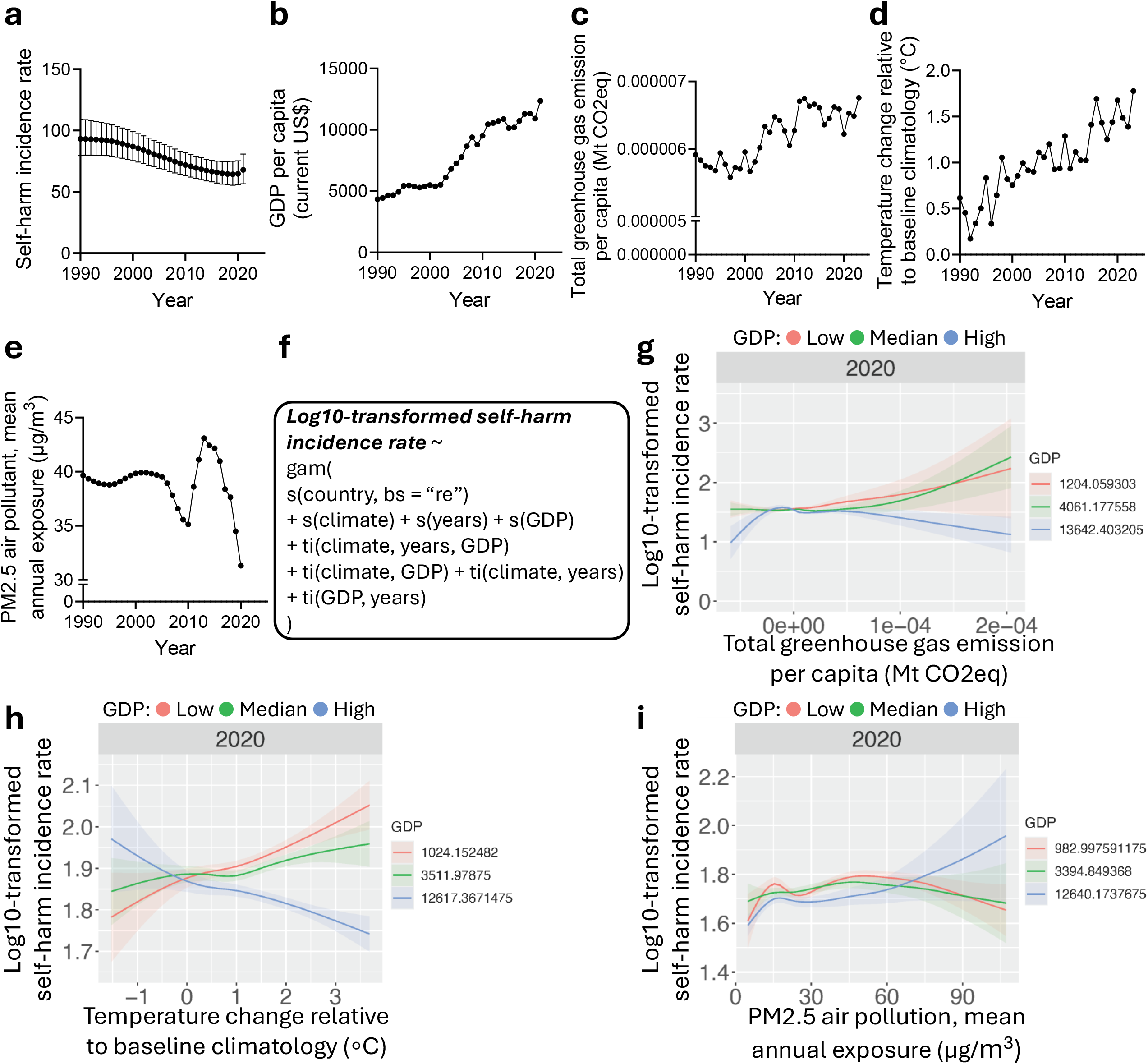
**a**. Global age-standardized self-harm incidence rate for both sexes as function of calendar year. **b**. Global gross domestic product (GDP) per capita as function of calendar year. **c**. Global total greenhouse gas emission per capita as function of calendar year. **d**. Global temperature changes relative to baseline climatology as function of calendar year. **e**. Global population-weighted particulate matter 2.5 (PM2.5) mean annual exposure as function of calendar year. **f**. Generative additive model (GAM) favoured in current analysis. **g**. Modelled effects of total greenhouse gas emission per capita on log10-transformed age-standardized self-harm incidence rate for both sexes globally at low (red), median (green), and high (blue) GDP. **h**. Modelled effects of temperature change relative to baseline climatology on log10-transformed age-standardized self-harm incidence rate for both sexes globally at low (red), median (green), and high (blue) GDP. **i**. Modelled effects of population-weighted PM2.5 mean annual exposure on log10-transformed age-standardized self-harm incidence rate for both sexes globally at low (red), median (green), and high (blue) GDP.

We have thus very recently conceptualized an analytical framework based on GAM for such purposes^9,10^. GAM is a state-of-the-art multi-dimensional analytical method for complex modelling, which captures the individual, additive and interactive impacts from various parameters (e.g., the climate change metrics) on outcomes of interest (e.g., self-harm incidence). It can also adjust for various confounders like socioeconomics (e.g., GDP), known to complicate self-harm and other suicidal behaviours^3^, in a similar manner to standard linear regression as applied in conventional epidemiological studies. GAM has been widely used by our team in a series of global health studies^13-17^. Details on our modelling are explained further in Methods and our recent publications^9,10^.

Here, we curated data for about 200 countries from 1990 to 2020. Harnessing GAM, we modelled self-harm incidence rate with major climate change-related parameters (i.e., GHG emission, temperature change, PM2.5 exposure and relative humidity) based on data spanning individual countries around the globe throughout years. Time (i.e., the calendar years that the data correspond to) and socioeconomic wealth (i.e., GDP) were adjusted in our models. Individual countries were modelled as random effects to partly cover the confounding effects from other inter-country differences like ethnicity, rural and urban distributions, etc., as is common practice in ecological analysis of this kind^13-17^. For all modelling, GAMs incorporating the main effects from climate change-related parameters (e.g., GHG emissions), time and GDP, plus their interactive effects were favoured (Figure 1f). With this approach, we demonstrated that climate change metrics, time and socioeconomic wealth were linked to self-harm incidence rates.

As in Figure 1g, GAM modelling results from 2020, the latest timepoint with complete data coverage, are shown as representative examples in marginal effect plots. Here, the log10-transformed age-standardized self-harm incidence rate is plotted as a function of the total GHG emission per capita and stratified by low (25%, red), median (50%, green), and high (75%, blue) GDP quantiles. For low and median GDP quantiles, GHG emission was linked to increased self-harm incidence rate, while in the high GDP quantile, milder effects were found (Table S2-3). Similarly, higher temperature change was linked to elevated self-harm incidence at the low and median GDP quantiles (Figure 1h, Table S4-5). In contrast, self-harm incidence rate was positively linked to PM2.5 exposure in the high GDP quantile and mildly fluctuated at the low and median quantiles (Figure 1i, Table S6-7). These results were consistent across earlier years with available data (e.g., 2010) (Figure S1a-c) and for sensitivity analyses, after excluding Greenland, the region with the highest self-harm incidence worldwide (Figure S1d-e), highlighting the robustness of our analyses. Country’s yearly average relative humidity was only mildly correlated with self-harm incidence as in Figure S1f.

Previous studies suggested that the impacts of climate change on suicidal behaviours vary for gender^4^ and age groups^18,19^, but detailed insights, particularly towards the potential interactions among these confounders, are lacking, and hence are a particular focus of this study. Globally, the incidence rates of self-harm, distinct from deaths due to suicide, is generally higher in females than males, but both peak in the 20-24-year age groups (Figure S2). We comprehensively modelled the influences of GHG emission, temperature change and PM2.5 exposure on self-harm incidence for both males and females across different age groups using the GAM approach described, revealing striking differences. Figure 2 shows representative results for young (20-24 years old), middle-aged (40-44 years old) and old adults (60-64 years old). Figure S3 presents the overall results for all age groups. At low and median GDP quantiles, GHG emission and temperature change generally correlated with more self-harm in males early in life (Figure 2a-b & Figure S3a-b). For these quantiles, PM2.5 was overall mildly associated with self-harm. In contrast, within the high GDP quantile, PM2.5 exposure was linked to more male self-harm early in life but more female self-harm later in life (Figure 2c & Figure S3c).

**Figure 2.**
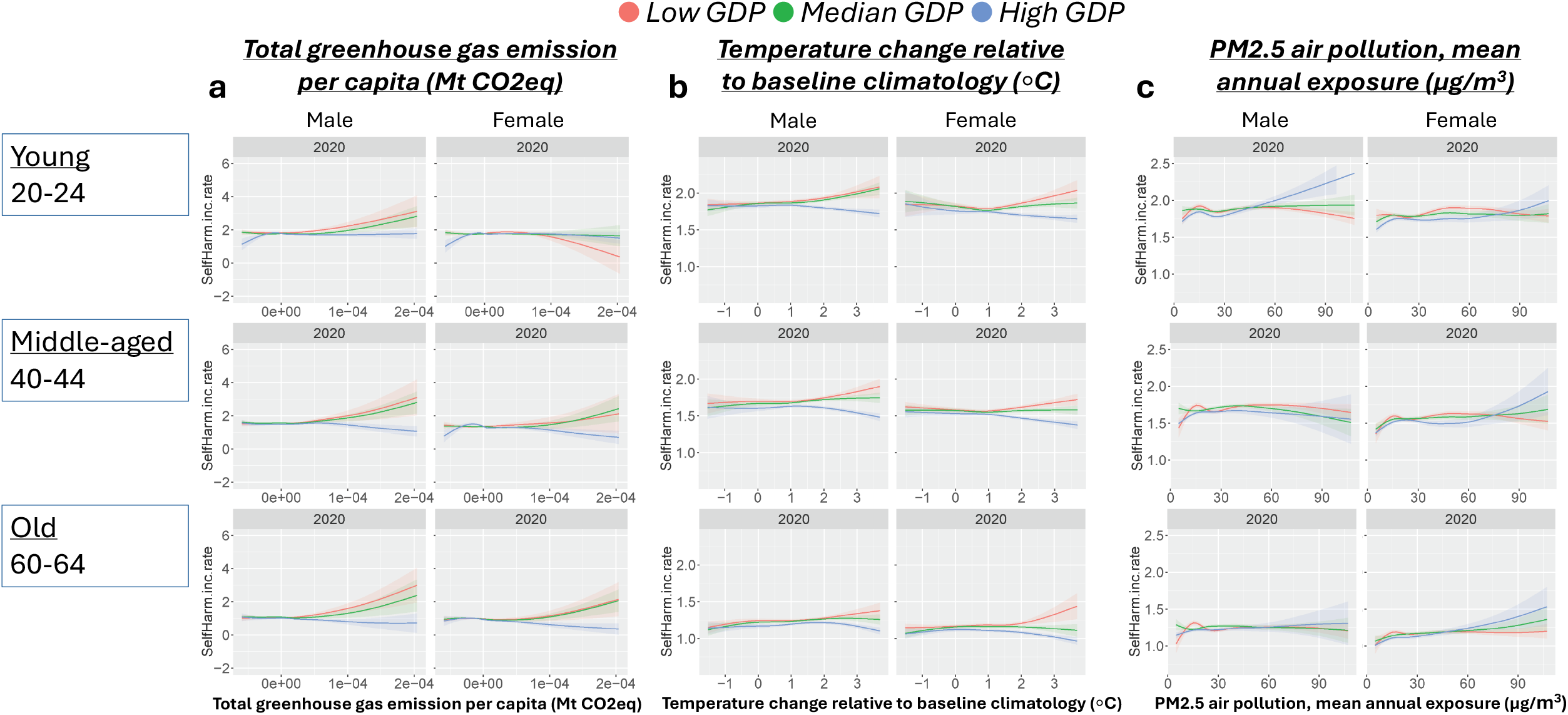
Modelled effects of total greenhouse gas emission per capita (**a**), temperature change relative to baseline climatology (**b**), and population-weighted PM2.5 mean annual exposure (**c**) on log10-transformed self-harm incidence rate in male (left columns) versus female (right columns) for representative young (20-24), middle-aged (40-44) and old (60-64) adults at low (red), median (green), and high (blue) GDP.

## Discussion

We systematically interrogated the associations between climate change and self-harm at an unprecedented global scale, covering over 200 countries worldwide across 30 years. Our GAM analyses provided compelling evidence that the major climate change-related parameters like GHG emissions and temperature change as well as air pollutant PM2.5 exposure, are generally associated with elevated self-harm incidence. In contrast, humidity seemed to play a modest role. We also revealed sophisticated interactions among gender, age and socioeconomic wealth (i.e., GDP) on the impacts of climate change on self-harm. Despite females generally having higher self-harm incidence rates than males, our analyses indicated that young males are more susceptible to climate change than young females, while the opposite was found later in life, depending on GDP. Our findings align with previous reports, which are mostly regional rather than global comparative studies and are generally based on smaller populations or are systematic reviews/meta-analyses^4,5,18-20^.

Self-harm represents a detrimental endpoint of a range of adverse social and population-based risk factors including common mental disorders, such as depression, anxiety and drug and alcohol use^3,12^. Future research interrogating how the spectrum of these risk factors, particularly the incidence of mental disorders among people, is affected by climate change might further complement our findings.

Analyses here cover major aspects of climate change like GHG emission, temperature change, as well as air pollutant PM2.5 exposure, an important contributor to climate change^11^, for which comprehensive global surveillance data is available. Analyses based on these yearly data reflected the chronic and slow-onset impacts from climate change and air pollution, complementing studies from us and others investigating the effects of sudden-onset climate events like acute heat waves, or other weather-related catastrophic events (e.g., cyclones, floods, etc.)^21-23^. Other aspects of climate change, like alterations in precipitation, drought, sea-level rise and El Niño/La Niña phenomena, might also play direct or indirect roles in mental health^3,22,24^. The analytical pipelines presented here can potentially extend to such studies in the future.

Our analyses are of correlative nature, leaving the underlying mechanisms unresolved. Climate change is known to directly contribute to anxiety, depression and grief^19^, particularly “eco-anxiety” regarding the consequences of climate change^25^. Climate change might also confer indirect impacts *via* disrupting livelihood, further increasing self-harm tendency, as exemplified by the associations between crop-damaging high temperatures and increased suicide rates in India^24^. Other potential influences may include biological effects on cellular and molecular levels, which might affect neurological and behavioural functions and development, resulting in increased rates of mental and behavioural disorders within the populations, particularly in young generations^23,26-28^. Our robust observations warrant future studies to demonstrate the causality and elucidate the underlying mechanisms, underpinning these associations. These endeavours might help to disentangle the complex gender-age interactions we observed, providing additional insights towards the gender paradox in suicide epidemiology^29-32^. Such research might also shed light on the stark contrasts found for countries with different socioeconomic statuses, as indicated by their varied GDP levels. In low and median GDP quantile countries, GHG emissions and temperature change, but not PM2.5 exposure, tend to associate with self-harm events. Contrarily, in high GDP countries, PM2.5 exposure positively correlates with self-harm. These prominent differences might stem from biological differences between gender over the lifespan such as neurodevelopmental and/or hormonal determinants^33-35^, as well as socioeconomic and cultural factors like livelihood and mood management^36-39^. These warrant further in-depth investigations and call for equity in global planetary health practice and policy making.

Global ecological analyses like ours are inevitably subject to various other confounders. In addition to the adjustments used in our GAMs (e.g., age, gender, socioeconomics) and the random effects to correct for inter-country differences, other factors like rates and types of substance use and geographical latitudes might also play a role^40^. Another limitation of our global analysis lies in intra-country data heterogeneity. Analyses here are based on country level data, but within the same country, influences from climate change might differ significantly geographically and vary within the population over time. Future studies with global regional data at higher resolutions are required.

Overall, we demonstrated for the first time the association between major climate change parameters and self-harm incidence rates on a global scale. Our findings underscore the significance of incorporating self-harm and mental health in future climate change and global health research. It will be useful in informing future public health practice and policy, leading to more nuanced and targeted global health campaigns to mitigate the impacts from climate change on human and planetary health.

## Methods

### Data collection and processing

Self-harm data was retrieved from the Global Burden of Disease database, and it is defined as *deliberate bodily damaged inflicted on oneself resulting in death or injury. ICD-9: E950-E959; ICD-10: X60-X64*.*9, X66-X84*.*9, Y87*.*0*. From World Bank (WB) databank (https://databank.worldbank.org/source/world-development-indicators), we obtained population, GDP (current US$ per capita), GHG emissions and PM2.5 exposure data. GDP is a close proxy of socioeconomic wealth, well-established in ecological studies^13-17^. It is considered in our model because wealth and income are known confounders for self-harm and other suicidal events^3^. Here, *total GHG emission data including land use, land use changes, and forestry* for each country from WB was divided by the population of the corresponding country at the corresponding timepoint to calculate the GHG emission per capita (Mt CO_2_eq per capita). PM2.5 exposure data was population-weighted (μg/m^3^). It referred to *the average level of exposure of a nation’s population to concentrations of suspended particles measuring less than 2*.*5 microns in aerodynamic diameter, which are capable of penetrating deep into the respiratory tract and causing severe health damage. Exposure is calculated by weighting mean annual concentrations of PM2*.*5 by population in both urban and rural areas*. Temperature change data (°C) was collected from Food and Agriculture Organization of the United Nations (https://www.fao.org/faostat/en/#data), calculated as *the temperature change on land with respect to baseline climatology, corresponding to the period 1951-1980*.

These data were compiled for analyses, and countries or timepoints without data documentation would be excluded. Overall, we curated several datasets covering close to 200 countries worldwide spanning 1990-2020 period for analyses.

### Generalized additive model (GAM)

Details of using generalized additive model (GAM) for global health analysis are described in our recent papers^9,10^. GAM excels in analyzing multiple parameters simultaneously, with consideration of their potential individual, additive and interactive effects, as well as their potential non-linear effects. This is achieved through the application of non-parametric smoothed functions, which are usually in forms of splines, providing flexible approaches to inspect the non-linear effects. GAM also adjusts for confounders in a similar manner to conventional epidemiological analyses. Therefore, GAM represents a powerful tool for global health analysis, where multiple factors are involved, requiring multi-dimensional thinking.

The GAMs we conceptualized^9,10^ leveraged various global parameters (i.e., global climate change parameters) and took into considerations of the potential confounders like time (i.e., the calendar years corresponding to the data) and socioeconomic wealth, reflected in GDP. The inter-country differences were modelled using random effects, a common practice in global ecological studies of this kind.

Here, an array of GAMs were run for log10-transformed self-harm incidence rates using different climate change parameters (i.e., GHG emission, temperature change and PM2.5 exposure) as predictors, together with time and GDP, using the *mgcv* package in its “*gam*” function in R. These GAMs considered the different combinations and interactions among all parameters. A null model where only the country-based random effects were considered was also included. Within these GAMs, individual parameters were modelled with smooth terms using the “s()” function, and their interactions were modelled with the “ti()” or “te()” functions, considering their different scales, like GHG emissions and GDP.

GAM results were evaluated based on Akaike information criterion (AIC) and the one with the lowest AIC was selected.

In our study, the model in Figure 1f was favoured based on AIC, where the main effects from climate change parameters, GDP and time/calendar year were modelled with smooth term “s()” and the interactive effects among them were modelled with tensor production interaction “ti()” function.

GAM results were visualized with *marginaleffects* package in R. As exemplified in Figure 1g, log10-transformed self-harm incidence rate (y-axis) was plotted as function of climate change parameter (x-axis, e.g., total greenhouse gas emission per capita), and the responses were stratified by GDP into low (red), median (green) and high (blue) quantiles.

## Figure legend

**Figure S1. a**. Modelled effects of total greenhouse gas emission per capita on log10-transformed age-standardized self-harm incidence rate for both sexes globally at low (red), median (green), and high (blue) GDP in 2010. **b**. Modelled effects of temperature change relative to baseline climatology on log10-transformed age-standardized self-harm incidence rate for both sexes globally at low (red), median (green), and high (blue) GDP in 2010. **c**. Modelled effects of population-weighted PM2.5 mean annual exposure on log10-transformed age-standardized self-harm incidence rate for both sexes globally at low (red), median (green), and high (blue) GDP in 2010. **d**. Modelled effects of temperature change relative to baseline climatology on log10-transformed age-standardized self-harm incidence rate for both sexes globally at low (red), median (green), and high (blue) GDP based on analysis excluding Greenland. **e**. Modelled effects of population-weighted PM2.5 mean annual exposure on log10-transformed age-standardized self-harm incidence rate for both sexes globally at low (red), median (green), and high (blue) GDP based on analysis excluding Greenland. **f**. Modelled effects of yearly average relative humidity on log10-transformed age-standardized self-harm incidence rate for both sexes globally at low (red), median (green), and high (blue) GDP.

**Figure S2**. Distribution of self-harm incidence rate for male versus female across age groups based on records from Global Burden of Disease.

**Figure S3**. Modelled effects of total greenhouse gas emission per capita (**a**), temperature change relative to baseline climatology (**b**), and population-weighted PM2.5 mean annual exposure (**c**) on log10-transformed self-harm incidence rate for male (left columns) versus female (right columns) across different age groups at low (red), median (green), and high (blue) GDP.

## Data availability

Data used in this study are all publicly available as described in Methods. Data for analysis in Figure 1 are available at: https://github.com/Nidane/ClimateChange-SelfHarm.

## Code availability

Codes used in this study are available at: https://github.com/Nidane/ClimateChange-SelfHarm.

## Acknowledgements

This project is supported by the Norman Ernest Bequest Fund.

## Author contributions

Concept and design: D.N. and R.N. Acquisition, analysis, and interpretation of data: D.N. and R.N. Drafting of the manuscript: D.N., I.B.H, M.H. and RN. Critical revision of the manuscript for important intellectual content: all authors. All authors read and approved the final manuscript.

