## Supplementary material for "Linking climate change to self-harm: A global study of over 200 countries from 1990 to 2020": Figure S1 & 2

*Time course analyses for earlier timepoint 2010*

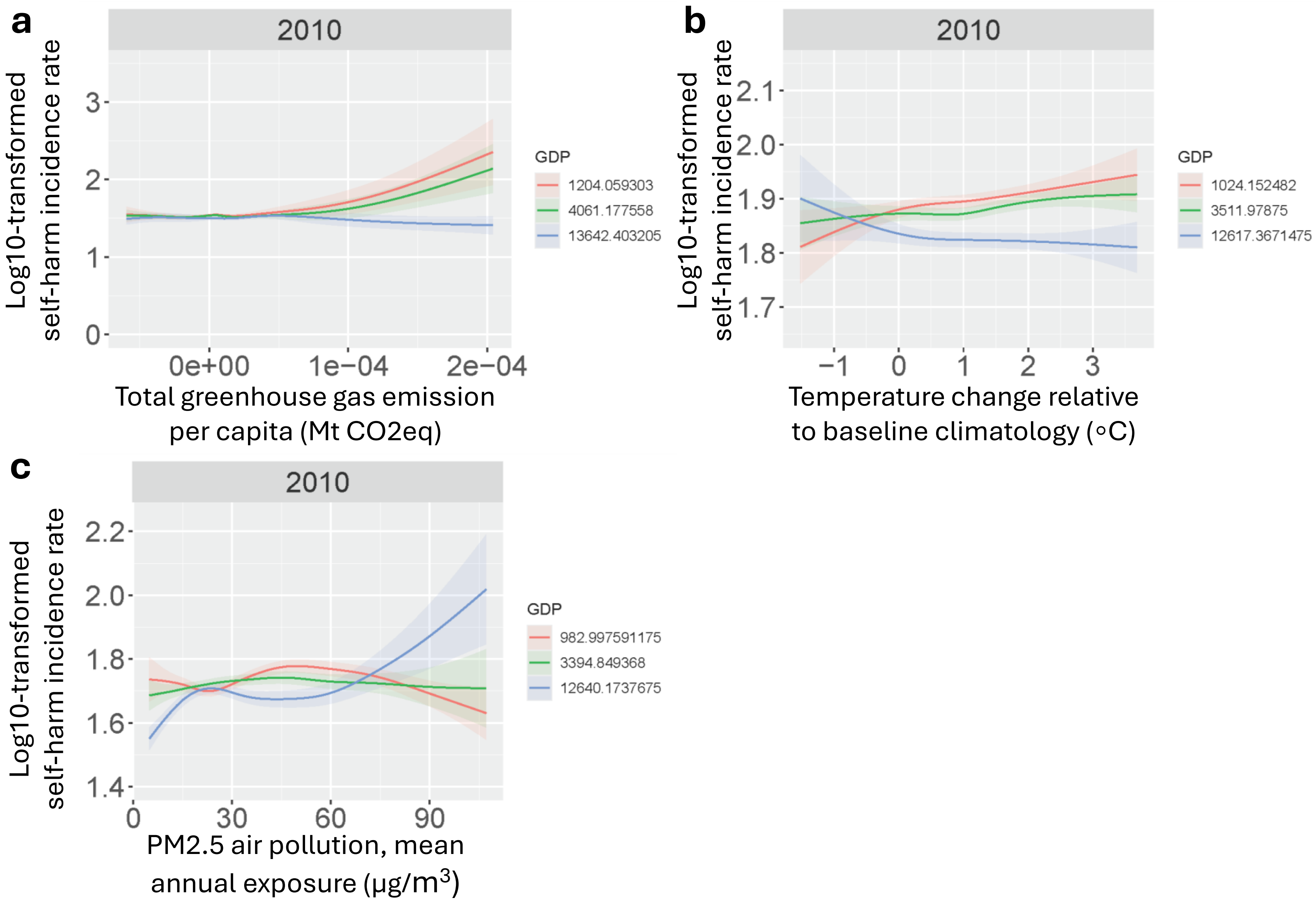


*Sensitivity analyses excluding Greenland*

*Note: Greenland does not have greenhouse gas emission record*

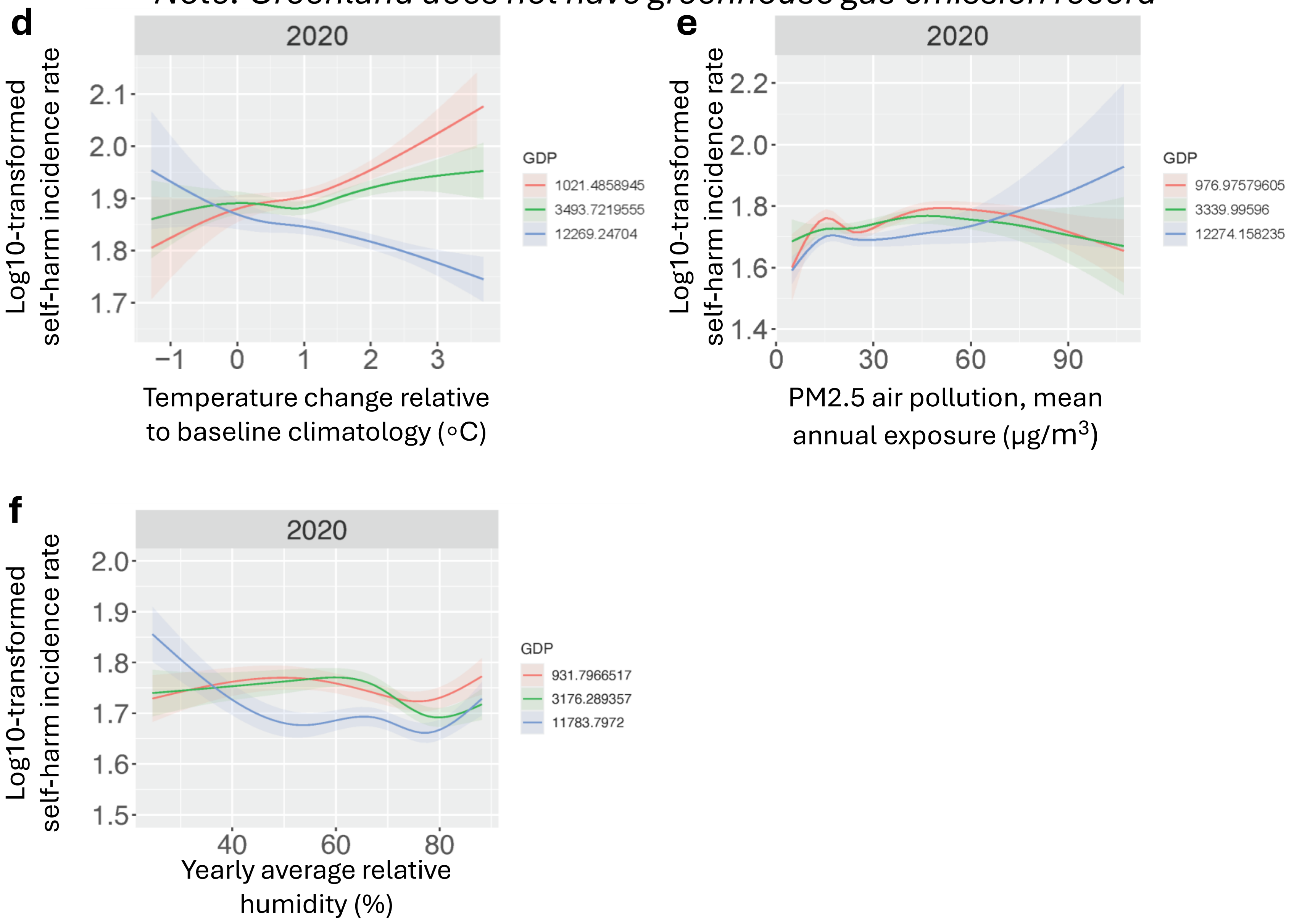

**Figure S2.**

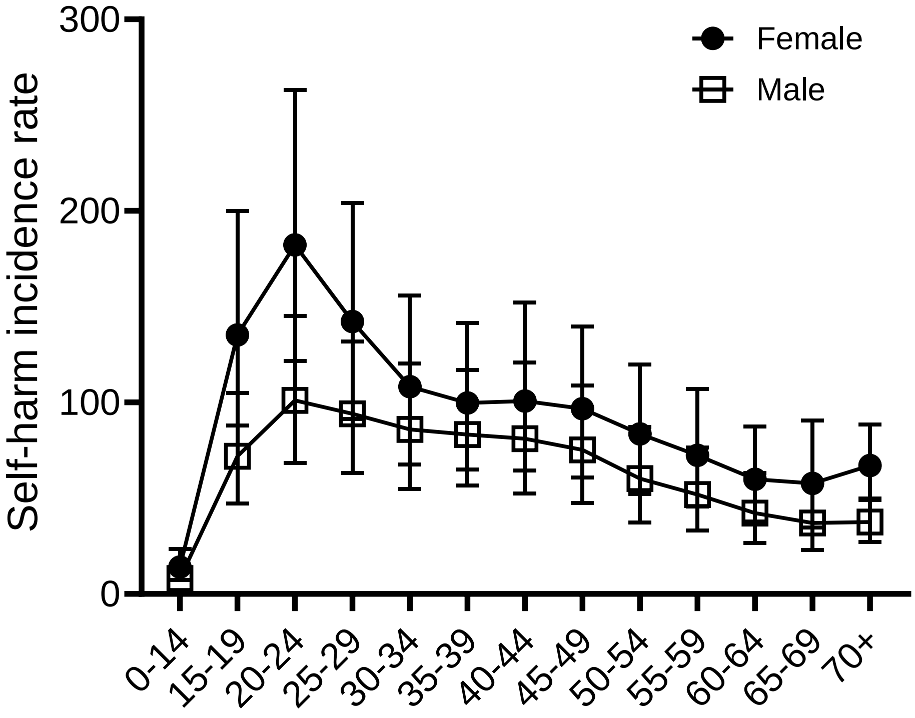
Female
Male
Self-harm incidence rate
300
200
100
0
0-14
15-19
20-24
25-29
30-34
35-39
40-44
45-49
50-54
55-59
60-64
65-69
70+
