## Supplementary figures and images for "Linking climate change to self-harm: A global study of over 200 countries from 1990 to 2020"

### Figure S3

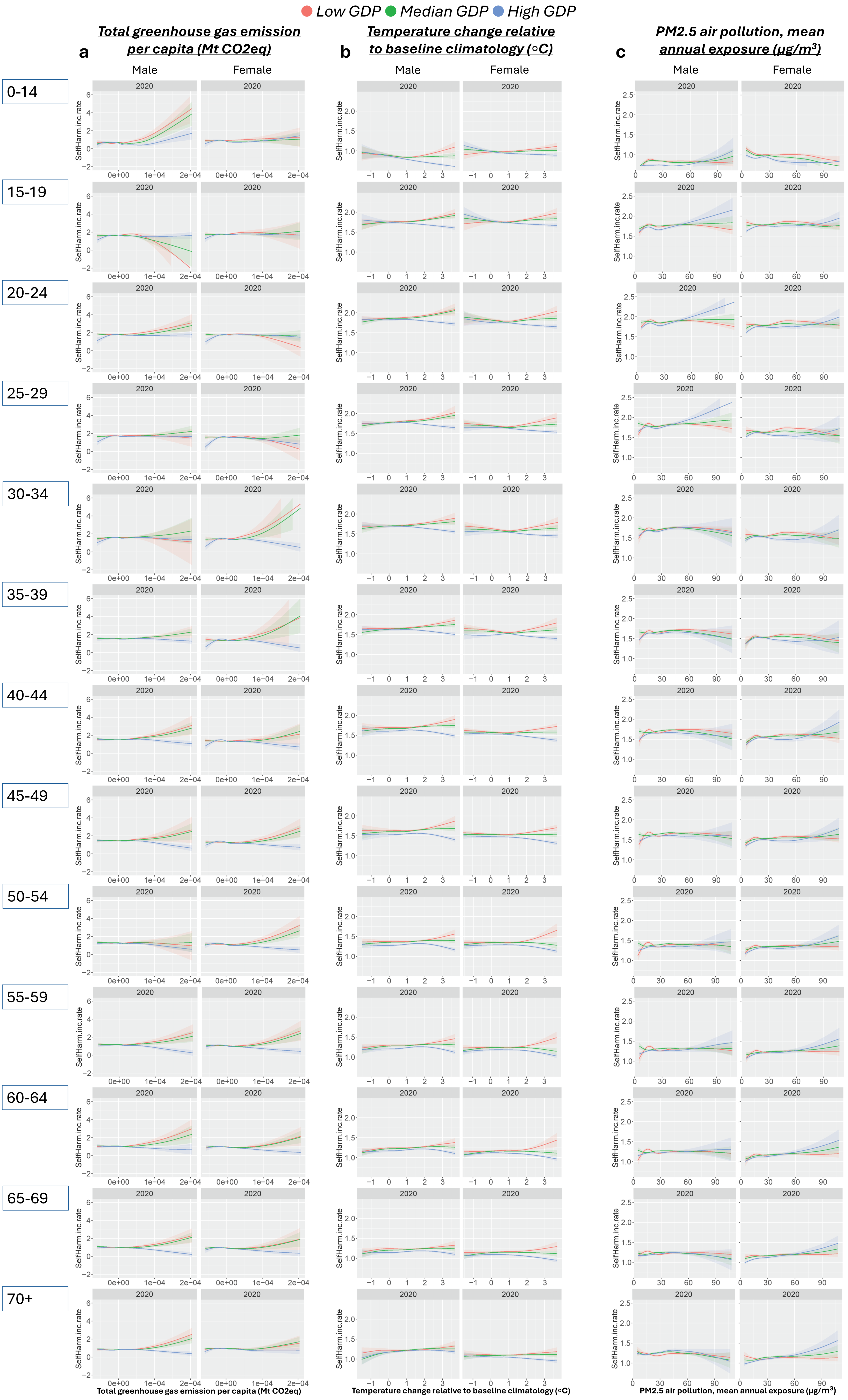
