## Supplementary Info for "Linking climate change to self-harm: A global study of over 200 countries from 1990 to 2020"

### Supplementary Tables

Table S2-7 are GAM estimates and statistical outputs. The model estimates and related standard errors (Std.Error) were shown for parametric terms. For non-parametric smooth terms, their estimated and reference degrees of freedom (i.e. edf, sumEDF, Ref.df) and their test statistics were shown.

**Table S1.** $R^2$ for correlations among global data.

| | **Self-harm** | **GDP** | **PM2.5** | **Temp** | **GHG** |
|---|---|---|---|---|---|
| **Self-harm** | 1.00 | 0.95 | 0.03 | 0.74 | 0.77 |
| **GDP** | - | 1.00 | 0.04 | 0.64 | 0.80 |
| **PM2.5** | - | - | 1.00 | 0.06 | 0.00 |
| **Temp** | - | - | - | 1.00 | 0.50 |
| **GHG** | - | - | - | - | 1.00 |

**Self-harm:** Self-harm age-standardized incidence rate for both sexes (new cases per 100000)
**GDP:** GDP per capita (current US$)
**PM2.5:** PM2.5 air pollution, mean annual exposure (µg/$m^3$)
**Temp:** Temperature change relative to baseline climatology (°C)
**GHG:** Total greenhouse gas (GHG) emissions per capita (Mt CO2eq)

**Table S2.** Relative fit of GAM analyzing the predictors for log10-transformed self-harm incidence rate globally (total greenhouse gas emission per capita). AIC = Akaike information criterion. sumEDF indicates the degrees of freedom of the models. Model 16 was selected as the best model because of the lowest AIC. (related to Figure 1g)

| GAM | AIC | sumEDF | Formula |
|---|---|---|---|
| 0 | -14557.42 | 174.84 | gam(log10-transformed self-harm incidence rate ~ 1 + s(Country, bs = "re")) |
| 1 | -15105.04 | 178.34 | gam(log10-transformed self-harm incidence rate ~ s(Country, bs = "re") + s(GDP)) |
| 2 | -14956.91 | 178.86 | gam(log10-transformed self-harm incidence rate ~ s(Country, bs = "re") + s(Year)) |
| 3 | -15231.06 | 186.49 | gam(log10-transformed self-harm incidence rate ~ s(Year) + s(GDP) + s(Country, bs = "re")) |
| 4 | -15346.1 | 193.38 | gam(log10-transformed self-harm incidence rate ~ te(Year, GDP) + s(Country, bs = "re")) |
| 5 | -14585.07 | 182.36 | gam(log10-transformed self-harm incidence rate ~ s(GHG.EMISSION) + s(Country, bs = "re")) |
| 6 | -14978.65 | 185.00 | gam(log10-transformed self-harm incidence rate ~ s(GHG.EMISSION) + s(Year) + s(Country, bs = "re")) |
| 7 | -15177.24 | 187.94 | gam(log10-transformed self-harm incidence rate ~ s(GHG.EMISSION) + s(GDP) + s(Country, bs = "re")) |
| 8 | -15242.28 | 191.88 | gam(log10-transformed self-harm incidence rate ~ s(GHG.EMISSION) + s(Year) + s(GDP) + s(Country, bs = "re")) |
| 9 | -15344.25 | 195.32 | gam(log10-transformed self-harm incidence rate ~ s(GHG.EMISSION) + te(Year, GDP) + s(Country, bs = "re")) |
| 10 | -15386.89 | 199.00 | gam(log10-transformed self-harm incidence rate ~ te(GHG.EMISSION, GDP) + s(Year) + s(Country, bs = "re")) |
| 11 | -15318.88 | 202.30 | gam(log10-transformed self-harm incidence rate ~ te(GHG.EMISSION, Year) + s(GDP) + s(Country, bs = "re")) |
| 12 | -15653.43 | 252.42 | gam(log10-transformed self-harm incidence rate ~ te(GHG.EMISSION, Year, GDP) + s(Country, bs = "re")) |
| 13 | -15065.14 | 192.29 | gam(log10-transformed self-harm incidence rate ~ te(GHG.EMISSION, Year) + s(Country, bs = "re")) |
| 14 | -15321.84 | 194.97 | gam(log10-transformed self-harm incidence rate ~ te(GHG.EMISSION, GDP) + s(Country, bs = "re")) |
| 15 | -15374.84 | 212.91 | gam(log10-transformed self-harm incidence rate ~ s(GHG.EMISSION) + s(Year) + s(GDP) + ti(GHG.EMISSION, Year, GDP) + s(Country, bs = "re"), data = complete |
| **16** | **-15708.42** | **233.82** | **gam(log10-transformed self-harm incidence rate ~ s(GHG.EMISSION) + s(Year) + s(GDP) + ti(GHG.EMISSION, Year, GDP) + ti(GHG.EMISSION, Year) + ti(GHG.EMISSION, GDP) + ti(GDP, Year) + s(Country, bs = "re"))** |
| 17 | -15515.49 | 215.45 | gam(log10-transformed self-harm incidence rate ~ s(GHG.EMISSION) + s(Year) + s(GDP) + ti(GHG.EMISSION, Year) + ti(GHG.EMISSION, GDP) + ti(GDP, Year) + s(Country, bs = "re")) |

| | | | |
|---|---|---|---|
| 18 | -15639.20 | 219.15 | gam(log10-transformed self-harm incidence rate ~ s(GHG.EMISSION) + s(Year) + s(GDP) + ti(GHG.EMISSION, Year, GDP) + ti(GHG.EMISSION, GDP) + ti(GDP, Year) + s(Country, bs = "re")) |
| 19 | -15646.26 | 217.28 | gam(log10-transformed self-harm incidence rate ~ s(GHG.EMISSION) + s(Year) + s(GDP) + ti(GHG.EMISSION, Year, GDP) + ti(GHG.EMISSION, Year) + ti(GDP, Year) + s(Country, bs = "re")) |
| 20 | -15634.85 | 221.01 | gam(log10-transformed self-harm incidence rate ~ s(GHG.EMISSION) + s(Year) + s(GDP) + ti(GHG.EMISSION, Year, GDP) + ti(GHG.EMISSION, Year) + ti(GHG.EMISSION, GDP) + s(Country, bs = "re")) |

**Table S3.** Estimated effects of total greenhouse gas (GHG) emission per capita, GDP per capita and year on log10-transformed self-harm incidence rate globally. (related to Figure 1g)

| **Parametric coefficients** | | | | |
|---|---|---|---|---|
| | **Estimate** | **Std.Error** | **t value** | **Pr(>\|t\|)** |
| **(Intercept)** | 1.646419 | 0.002329 | 707.1 | <2e-16 |
| **Approximate significance of smooth terms** | | | | |
| | **edf** | **Ref.df** | **F** | **P value** |
| **s(GHG.EMISSION)** | 6.743 | 7.941 | 1.756 | 0.0988 |
| **s(Year)** | 4.866 | 5.956 | 16.199 | <2e-16 |
| **s(GDP)** | 8.005 | 8.742 | 9.376 | <2e-16 |
| **ti(GHG.EMISSION, Year, GDP)** | 17.527 | 19.358 | 11.234 | <2e-16 |
| **ti(GHG.EMISSION, Year)** | 4.000 | 4.000 | 14.776 | <2e-16 |
| **ti(GHG.EMISSION, GDP)** | 8.865 | 9.670 | 6.051 | <2e-16 |
| **ti(Year, GDP)** | 8.817 | 10.280 | 7.600 | <2e-16 |
| **s(Country)** | 174.006 | 175.000 | 1605.212 | <2e-16 |
| R-sq.(adj) = 0.992 Deviance explained = 99.2% | | | | |
| GCV = 0.00080387 Scale est. = 0.00075251 n = 3660 | | | | |

**Table S4.** Relative fit of GAM analyzing the predictors for log10-transformed self-harm incidence rate globally (temperature change relative to baseline climatology). AIC = Akaike information criterion. sumEDF indicates the degrees of freedom of the models. Model 16 was selected as the best model because of the lowest AIC. (related to Figure 1h)

| GAM | AIC | sumEDF | Formula |
|---|---|---|---|
| 0 | -19582.21 | 195.77 | gam(log10-transformed self-harm incidence rate ~ 1 + s(Country, bs = "re")) |
| 1 | -20294.13 | 204.61 | gam(log10-transformed self-harm incidence rate ~ s(Country, bs = "re") + s(GDP)) |
| 2 | -19935.22 | 201.68 | gam(log10-transformed self-harm incidence rate ~ s(Country, bs = "re") + s(Year)) |
| 3 | -20357.67 | 211.64 | gam(log10-transformed self-harm incidence rate ~ s(Year) + s(GDP) + s(Country, bs = "re")) |
| 4 | -20518.72 | 218.18 | gam(log10-transformed self-harm incidence rate ~ te(Year, GDP) + s(Country, bs = "re")) |
| 5 | -19703.61 | 201.53 | gam(log10-transformed self-harm incidence rate ~ s(TEMPCHANGE) + s(Country, bs = "re")) |
| 6 | -19978.92 | 207.08 | gam(log10-transformed self-harm incidence rate ~ s(TEMPCHANGE) + s(Year) + s(Country, bs = "re")) |
| 7 | -20317.47 | 209.92 | gam(log10-transformed self-harm incidence rate ~ s(TEMPCHANGE) + s(GDP) + s(Country, bs = "re")) |
| 8 | -20383.29 | 217.10 | gam(log10-transformed self-harm incidence rate ~ s(TEMPCHANGE) + s(Year) + s(GDP) + s(Country, bs = "re")) |
| 9 | -20532.60 | 223.60 | gam(log10-transformed self-harm incidence rate ~ s(TEMPCHANGE) + te(Year, GDP) + s(Country, bs = "re")) |
| 10 | -20456.89 | 222.62 | gam(log10-transformed self-harm incidence rate ~ te(TEMPCHANGE, GDP) + s(Year) + s(Country, bs = "re")) |
| 11 | -20395.63 | 222.50 | gam(log10-transformed self-harm incidence rate ~ te(TEMPCHANGE, Year) + s(GDP) + s(Country, bs = "re")) |
| 12 | -20697.49 | 269.46 | gam(log10-transformed self-harm incidence rate ~ te(TEMPCHANGE, Year, GDP) + s(Country, bs = "re")) |
| 13 | -20019.53 | 214.24 | gam(log10-transformed self-harm incidence rate ~ te(TEMPCHANGE, Year) + s(Country, bs = "re")) |
| 14 | -20391.18 | 215.51 | gam(log10-transformed self-harm incidence rate ~ te(TEMPCHANGE, GDP) + s(Country, bs = "re")) |
| 15 | -20646.97 | 237.61 | gam(log10-transformed self-harm incidence rate ~ s(TEMPCHANGE) + s(Year) + s(GDP) + ti(TEMPCHANGE, Year, GDP) + s(Country, bs = "re"), data = complete |
| **16** | **-20764.19** | **249.59** | **gam(log10-transformed self-harm incidence rate ~ s(TEMPCHANGE) + s(Year) + s(GDP) + ti(TEMPCHANGE, Year, GDP) + ti(TEMPCHANGE, Year) + ti(TEMPCHANGE, GDP) + ti(GDP, Year) + s(Country, bs = "re"))** |
| 17 | -20686.44 | 243.74 | gam(log10-transformed self-harm incidence rate ~ s(TEMPCHANGE) + s(Year) + s(GDP) + ti(TEMPCHANGE, Year) + ti(TEMPCHANGE, GDP) + ti(GDP, Year) + s(Country, bs = "re")) |

| | | | |
|---|---|---|---|
| 18 | -20763.32 | 248.13 | gam(log10-transformed self-harm incidence rate ~ s(TEMPCHANGE) + s(Year) + s(GDP) + ti(TEMPCHANGE, Year, GDP) + ti(TEMPCHANGE, GDP) + ti(GDP, Year) + s(Country, bs = "re")) |
| 19 | -20759.4 | 240.51 | gam(log10-transformed self-harm incidence rate ~ s(TEMPCHANGE) + s(Year) + s(GDP) + ti(TEMPCHANGE, Year, GDP) + ti(TEMPCHANGE, Year) + ti(GDP, Year) + s(Country, bs = "re")) |
| 20 | -20677.04 | 265.41 | gam(log10-transformed self-harm incidence rate ~ s(TEMPCHANGE) + s(Year) + s(GDP) + ti(TEMPCHANGE, Year, GDP) + ti(TEMPCHANGE, Year) + ti(TEMPCHANGE, GDP) + s(Country, bs = "re")) |

**Table S5.** Estimated effects of temperature change relative to baseline climatology, GDP per capita and year on log10-transformed self-harm incidence rate globally. (related to Figure 1h)

| **Parametric coefficients** | | | | |
|---|---|---|---|---|
| | **Estimate** | **Std.Error** | **t value** | **Pr(>\|t\|)** |
| **(Intercept)** | 1.667 | 0.543 | 3.071 | 0.00214 |
| **Approximate significance of smooth terms** | | | | |
| | **edf** | **Ref.df** | **F** | **P value** |
| **s(TEMPCHANGE)** | 1.635 | 2.092 | 2.880 | 0.0604 |
| **s(Year)** | 6.696 | 7.825 | 7.183 | <2e-16 |
| **s(GDP)** | 8.768 | 8.978 | 20.117 | <2e-16 |
| **ti(TEMPCHANGE, Year, GDP)** | 11.652 | 12.797 | 8.442 | <2e-16 |
| **ti(TEMPCHANGE, Year)** | 2.207 | 3.033 | 1.422 | 0.2323 |
| **ti(TEMPCHANGE, GDP)** | 9.338 | 10.412 | 1.694 | 0.0786 |
| **ti(Year, GDP)** | 13.296 | 14.475 | 9.008 | <2e-16 |
| **s(Country)** | 195.000 | 196.000 | 1269.356 | <2e-16 |
| R-sq.(adj) = 0.982 Deviance explained = 98.3% | | | | |
| GCV = 0.0018607 Scale est. = 0.00017836 n = 6018 | | | | |

**Table S6.** Relative fit of GAM analyzing the predictors for log10-transformed self-harm incidence rate globally (PM2.5 mean annual exposure). AIC = Akaike information criterion. sumEDF indicates the degrees of freedom of the models. Model 16 was selected as the best model because of the lowest AIC. (related to Figure 1i)

| GAM | AIC | sumEDF | Formula |
|---|---|---|---|
| 0 | -20135.66 | 198.80 | gam(log10-transformed self-harm incidence rate ~ 1 + s(Country, bs = "re")) |
| 1 | -20815.68 | 207.67 | gam(log10-transformed self-harm incidence rate ~ s(Country, bs = "re") + s(GDP)) |
| 2 | -20507.09 | 203.71 | gam(log10-transformed self-harm incidence rate ~ s(Country, bs = "re") + s(Year)) |
| 3 | -20864.59 | 213.17 | gam(log10-transformed self-harm incidence rate ~ s(Year) + s(GDP) + s(Country, bs = "re")) |
| 4 | -21008.88 | 221.05 | gam(log10-transformed self-harm incidence rate ~ te(Year, GDP) + s(Country, bs = "re")) |
| 5 | -20448.10 | 200.61 | gam(log10-transformed self-harm incidence rate ~ s(PM2.5) + s(Country, bs = "re")) |
| 6 | -20884.55 | 212.04 | gam(log10-transformed self-harm incidence rate ~ s(PM2.5) + s(Year) + s(Country, bs = "re")) |
| 7 | -20983.55 | 212.23 | gam(log10-transformed self-harm incidence rate ~ s(PM2.5) + s(GDP) + s(Country, bs = "re")) |
| 8 | -20985.65 | 216.42 | gam(log10-transformed self-harm incidence rate ~ s(PM2.5) + s(Year) + s(GDP) + s(Country, bs = "re")) |
| 9 | -21145.53 | 225.47 | gam(log10-transformed self-harm incidence rate ~ s(PM2.5) + te(Year, GDP) + s(Country, bs = "re")) |
| 10 | -21312.38 | 227.40 | gam(log10-transformed self-harm incidence rate ~ te(PM2.5, GDP) + s(Year) + s(Country, bs = "re")) |
| 11 | -21190.79 | 226.54 | gam(log10-transformed self-harm incidence rate ~ te(PM2.5, Year) + s(GDP) + s(Country, bs = "re")) |
| 12 | -21783.36 | 299.76 | gam(log10-transformed self-harm incidence rate ~ te(PM2.5, Year, GDP) + s(Country, bs = "re")) |
| 13 | -21087.55 | 218.81 | gam(log10-transformed self-harm incidence rate ~ te(PM2.5, Year) + s(Country, bs = "re")) |
| 14 | -21258.99 | 221.72 | gam(log10-transformed self-harm incidence rate ~ te(PM2.5, GDP) + s(Country, bs = "re")) |
| 15 | -21311.9 | 263.43 | gam(log10-transformed self-harm incidence rate ~ s(PM2.5) + s(Year) + s(GDP) + ti(PM2.5, Year, GDP) + s(Country, bs = "re"), data = complete |
| **16** | **-21841.08** | **305.11** | **gam(log10-transformed self-harm incidence rate ~ s(PM2.5) + s(Year) + s(GDP) + ti(PM2.5, Year, GDP) + ti(PM2.5, Year) + ti(PM2.5, GDP) + ti(GDP, Year) + s(Country, bs = "re"))** |
| 17 | -21724.29 | 256.44 | gam(log10-transformed self-harm incidence rate ~ s(PM2.5) + s(Year) + s(GDP) + ti(PM2.5, Year) + ti(PM2.5, GDP) + ti(GDP, Year) + s(Country, bs = "re")) |

| | | | |
|---|---|---|---|
| 18 | -21743.06 | 298.52 | gam(log10-transformed self-harm incidence rate ~ s(PM2.5) + s(Year) + s(GDP) + ti(PM2.5, Year, GDP) + ti(PM2.5, GDP) + ti(GDP, Year) + s(Country, bs = "re")) |
| 19 | -21515.79 | 276.52 | gam(log10-transformed self-harm incidence rate ~ s(PM2.5) + s(Year) + s(GDP) + ti(PM2.5, Year, GDP) + ti(PM2.5, Year) + ti(GDP, Year) + s(Country, bs = "re")) |
| 20 | -21767.8 | 295.5 | gam(log10-transformed self-harm incidence rate ~ s(PM2.5) + s(Year) + s(GDP) + ti(PM2.5, Year, GDP) + ti(PM2.5, Year) + ti(PM2.5, GDP) + s(Country, bs = "re")) |

**Table S7.** Estimated effects of PM2.5 mean annual exposure, GDP per capita and year on log10-transformed self-harm incidence rate globally. (related to Figure 1i)

| **Parametric coefficients** | | | | |
|---|---|---|---|---|
| | **Estimate** | **Std.Error** | **t value** | **Pr(>\|t\|)** |
| **(Intercept)** | 1.6778 | 0.1836 | 9.137 | <2e-16 |
| **Approximate significance of smooth terms** | | | | |
| | **edf** | **Ref.df** | **F** | **P value** |
| **s(PM2.5)** | 7.750 | 8.622 | 7.204 | <2e-16 |
| **s(Year)** | 5.554 | 6.801 | 6.654 | <2e-16 |
| **s(GDP)** | 8.665 | 8.958 | 17.696 | <2e-16 |
| **ti(PM2.5, Year, GDP)** | 49.798 | 53.180 | 3.582 | <2e-16 |
| **ti(PM2.5, Year)** | 9.888 | 11.156 | 12.977 | <2e-16 |
| **ti(PM2.5, GDP)** | 15.222 | 15.570 | 22.144 | <2e-16 |
| **ti(Year, GDP)** | 9.234 | 10.772 | 8.207 | <2e-16 |
| **s(Country)** | 197.997 | 199.000 | 1452.595 | <2e-16 |
| R-sq.(adj) = 0.985 Deviance explained = 98.6% | | | | |
| GCV = 0.0015723 Scale est. = 0.0014928 n = 6034 | | | | |
